# Regulatory genomic circuitry of brain age by integrative functional genomic analyses

**DOI:** 10.1101/2025.02.23.25322727

**Authors:** Xingzhong Zhao, Anyi Yang, Jing Ding, Yucheng T. Yang, Xing-Ming Zhao

## Abstract

Brain age gap (BAG) is a valuable biomarker for evaluating brain healthy status and detecting age-associated cognitive degeneration. However, the genetic architecture of BAG and the underlying mechanisms are poorly understood. Here, we estimate brain age from magnetic resonance imaging with improved accuracy using our proposed adversarial convolution network (ACN), followed by applying the ACN model to an elder cohort from UK Biobank. The genetic heritability of BAG is significantly enriched in the regulatory regions and implicated in glial cells. We prioritize a set of BAG-associated genes, and further characterize their expression patterns across brain cell types and regions. Two BAG-associated genes, *RUNX2* and *KLF3*, are found as associated with epigenetic clock and diverse aging-related biological pathways. Finally, two BAG-associated hub transcription factors, *KLF3* and *SOX10*, are identified as regulators of pleiotropic risk genes from diverse brain disorders. Altogether, we improve the estimation of BAG, and identify BAG-associated genes and regulatory networks that are implicated in brain disorders.

## Introduction

As a complex biological phenomenon, brain aging is hallmarked by the progressive accrual of molecular and cellular damage over an individual’s lifetime and instigates the emergence of chronic brain diseases and cognitive decline[1–3]. Unveiling the molecular mechanisms underpinning brain aging is critical for preserving brain health[2]. Structural magnetic resonance imaging (MRI) has proven to be a valuable technology for quantitatively gauging the aging status of the brain[4], while machine learning and deep learning methodologies have been utilized to predict “brain age” by extracting aging-related features from structured MRI images[5–7]. The difference between brain age and chronological age, termed the "brain age gap" (BAG), could be used to discern whether an individual’s brain appears elder or younger than that of an age-matched healthy one. BAG has been proven as a novel biomarker for evaluating the degree of brain aging[6].

Previous studies have established the associations between BAG and cognitive impairments, as well as psychiatric disorders[8–10]. For example, patients diagnosed with Alzheimer’s disease (AD) show notably larger BAG scores than healthy participants, consistent with the observation that AD can expedite the process of brain aging[11, 12]. BAG has been proven to be heritable with a heritability score of ∼0.2[13, 14]. BAG also shows polygenic overlap with diverse psychiatric disorders, such as schizophrenia and bipolar disorder[13, 14]. BAG provides a potent medium to bridge brain aging and brain disorders, making it necessary to systematically explore the shared genetic architecture and molecular mechanisms between brain aging and brain disorders.

Brain structure has been shown as accompanied by intricate modifications in cellular senescence and molecular pathways during the aging process[15, 16]. Transcriptomic data from brain cells and tissues at different developmental stages can be employed to identify the genes associated with brain aging by comparing gene expression profiles across different stages[13, 17, 18]. For example, Buckley *et al.* revealed that the transcriptomic signatures from microglia can accurately estimate the degree of brain aging[19]. Kang *et al.* showed that the decline in synaptic functions during brain aging can be triggered by the increased expression of *REST* and decreased expression of *TP73*[18]. However, the association between the molecular signatures and the dynamics of brain structures during brain aging remains unclear. To which degree BAG can represent the process of brain aging and the association of BAG with the molecular changes during brain aging need to be clarified. The availability of large amounts of brain imaging data, as well as functional genomic data from human brain provides an excellent opportunity to explore these questions.

In this study, we characterized the genetic architecture of BAG by developing an adversarial convolution network (ACN) with improved generality and accuracy in brain age prediction, and then applying the ACN model to an elder cohort from UK Biobank (UKB). We further explored the genetic basis of BAG and the underlying regulatory mechanisms of BAG-associated genetic variants and genes with functional genomics analysis. The genetic heritability of BAG was significantly enriched in the regulatory regions and implicated in glial cells. We identified nine novel significant variants and 44 novel BAG-associated genes. We showed that the BAG-associated genes exhibited elevated expression levels in aging-related brain regions and cell types. Interestingly, BAG shows concordant patterns with epigenetic clock and aging-related biological pathways. Furthermore, we identified two BAG-associated hub transcription factors (TFs), *KLF3* and *SOX10* regulating pleiotropic risk genes of brain disorders. The regulatory target genes of *KLF3* and *SOX10* are enriched with neuroinflammation-related functions. To summarize, our study improved the methodology of BAG estimation, and revealed the genetic architecture and molecular mechanisms of BAG by incorporating diverse functional genomic data from the human brain.

## Results

### Predicting brain age based on MRI with improved accuracy

The heterogeneity of medical images often hampers the generalization capability of machine learning and deep learning models, posing challenges when applied to new datasets. To overcome these limitations, we developed an adversarial convolution network (ACN) (Figure S1), which was based on deep learning for accurately predicting brain age. Briefly, our model utilized a domain-adversarial training strategy[20], wherein we considered irrelevant factors (e.g., gender and data sites) as unique domains in mixed modules and implemented dynamic restrictions on these modules during the training process (**Figure 1a** and Figure S1; Methods). To improve the generality and robustness of our model in predicting BAG, we performed the model training and evaluation on the MRI datasets from 2,011 healthy participants with ages ranging from 4 to 95 years, which were collected from five independent datasets (**Figure 1b** and Table S1; Methods), including the Southwest University Adult Lifespan Dataset (SALD)[21], Citigroup Biomedical Imaging Center (CBIC)[22], Australian Imaging Biomarkers and Lifestyle Study of Ageing (AIBL)[23], IXI, and Open Access Series of Imaging Studies (OASIS1)[24]. We compared the performance of our ACN model with other models (**Table 1** and Table S2), including LASSO regression, Elastic network, ridge regression, Gaussian process regression (GPR), support vector regression (SVR), 3D ResNet[14], SFCN[7], as well as the ACN-based baseline models. We found that the ACN model showed superior performance over other models in terms of mean absolute error (MAE) (**Table 1**).

**Figure 1.**
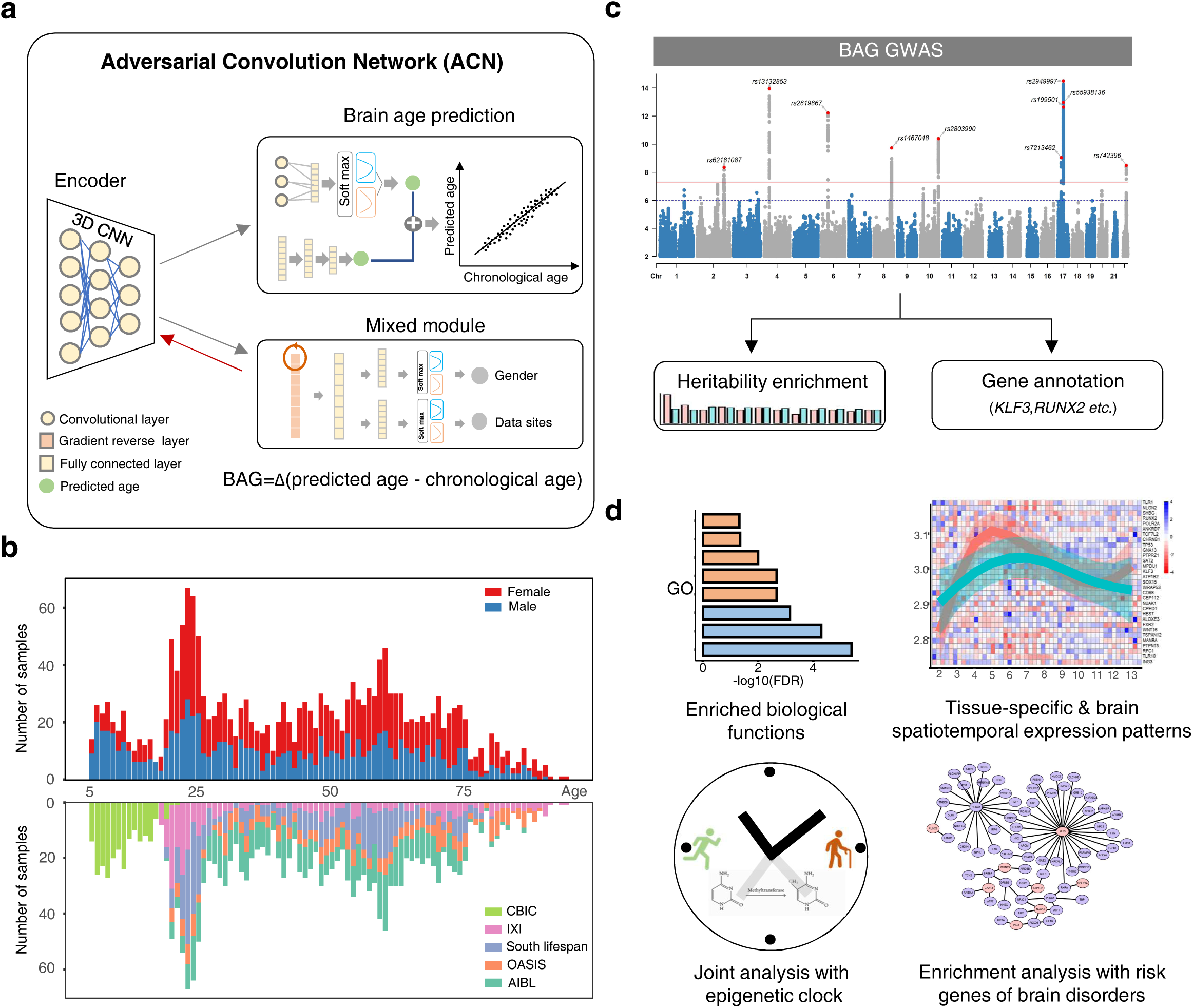
The workflow for the prediction of BAG and BAG GWAS, followed by analyses revealing the biological implications and relationships with brain disorders for BAG. (**a**) The ACN framework for the prediction of brain age (Methods). (**b**) The distribution of age and gender for the participants from five independent datasets including CBIC, IXI, SALD, OASIS and AIBL. (**c**) GWAS analysis of BAG and post-GWAS analysis. (**d**) Characterizing the enriched biological functions, the expression dynamics, the relationships with epigenetic clock and brain disorders of BAG-associated genes.

Next, we sought to validate the efficiency and robustness of ACN. We first explored the capabilities of ACN by restraining encoders from acquiring confounding factors, including gender and data sites. We extracted stratified features (i.e., features after encoding) of training data (Figure S1), and visualized their distributions with respect to gender and data sites using *t*-SNE (Figure S2 and S3). We found that ACN enhanced the generalization performance of encoders by effectively combining features from different genders and data sites. Secondly, we examined whether the stratified features were associated with brain aging. To this end, we used our pre-trained ACN model to extract the stratified features for the participants from Alzheimer’s Disease Neuroimaging Initiative (ADNI)[25] (N=437, including 150 AD patients, ages ranging from 55 to 91 years), in which the individuals were diagnosed with a typical brain aging-related disorder (Methods). Subsequently, we utilized a Support Vector Machine (SVM) classifier for diagnosing AD based on the stratified features. The SVM classifier achieved high accuracy (0.79 ± 0.28, with 5-fold cross-validation) in distinguishing the AD patients based on the stratified features (Figure S4), suggesting that the stratified features are biologically meaningful and the brain imaging features derived from AD patients are associated with brain aging[26].

### Identification of novel BAG-associated genetic variants

Knowledge of the genetic basis of BAG has markedly improved in recent years[13, 27, 28]. However, identifying novel genetic variants and genes associated with BAG in sizable studies remains challenging. The UKB dataset is ideal for the genetic analysis of BAG because it provides MRI image data and genotype data from the same individual and in a large sample size[29]. However, we observed an obvious age distribution difference between the UKB dataset (ages ranging from 45 to 85 years) and our training set (ages ranging from 5 to 94 years), which may bias the results of model prediction. To this end, we adopted a transfer learning approach to fine-tune the pre-trained ACN model, and then applied the fine-tuned model to the elder cohort from UKB (Methods).

In our study, we performed the GWAS analysis on BAG based on a larger sample size (N=35,702) than previous studies[13, 27, 28, 30]. Briefly, we estimated the BAG for 35,702 participants using the ACN model (MAE = 2.70; Figure S5), and then performed GWAS for BAG using GCTA[31] (Methods). In total, 3,868 genetic variants that exceeded genome-wide significance (*P* < 5e-08) were identified in our BAG GWAS (Table S3). Following linkage disequilibrium (LD)–based clumping (LD *r*^2^ < 0.1), we identified ten lead variants, among which nine were novel and one (rs13132853) has been reported as the lead SNP by recent study[28] (**Figure 2a**, Figure S6 and Table S4). In addition, five out of the nine novel lead SNPs exceeded genome-wide significance (*P* < 5e-08) in a recent study[28] (Table S5).

**Figure 2.**
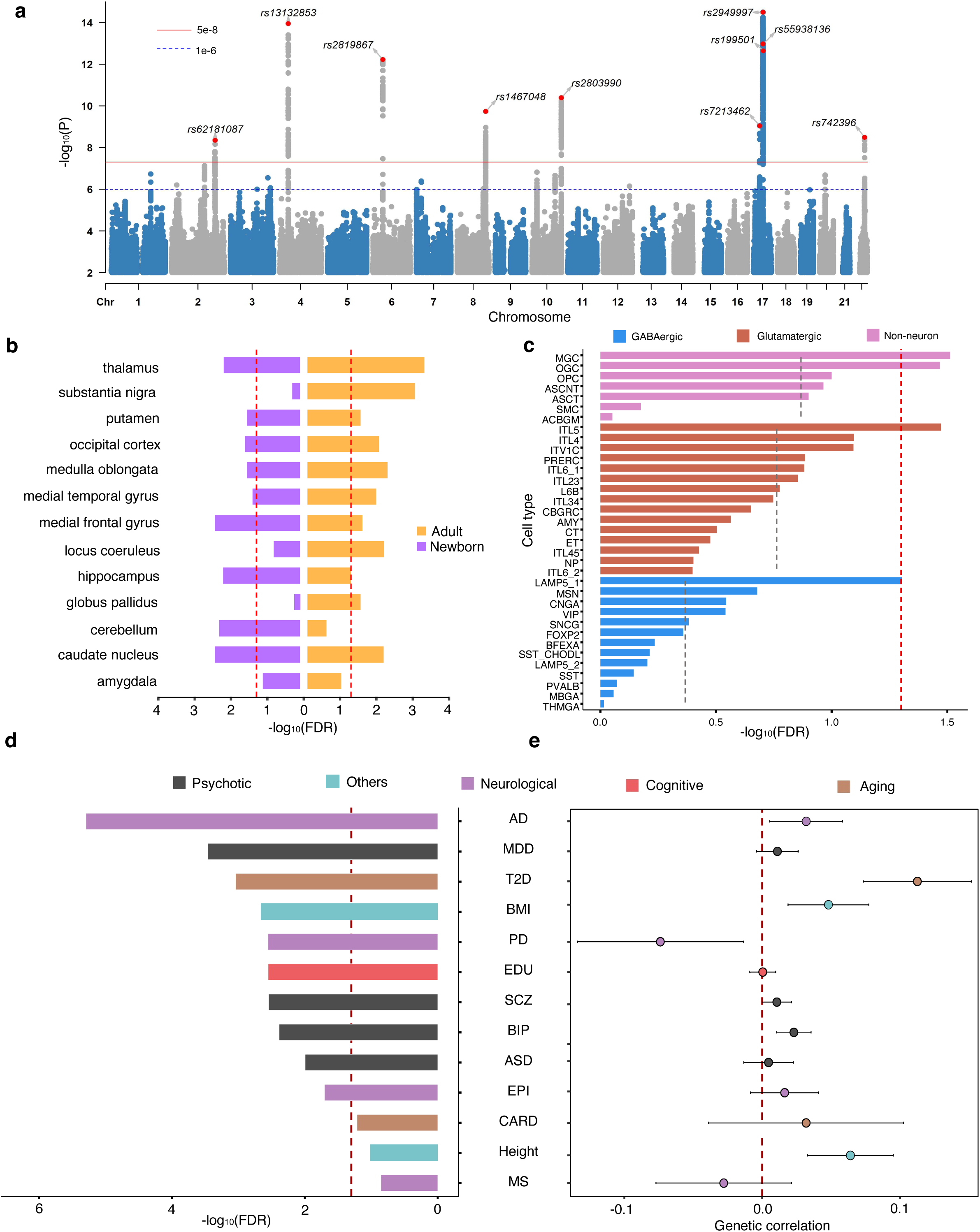
GWAS analysis of BAG. (**a**) GWAS discovery of BAG (Methods). The red line indicates GWAS summary statistics *P* threshold at 5e-08. (**b**) Partitioned heritability enrichment in active cCREs from brain regions. The red dashed line corresponds to an *FDR* threshold at the 5% level. (**c**) Partitioned heritability enrichment in active cCREs from neural cell types. The red dashed line corresponds to an *FDR* threshold at the 5% level. The detailed names of the cell types are provided in Table S6. (**d**) The associations between BAG and the polygenic scores of the 13 complex traits estimated using a linear regression model (Methods). The red dashed line corresponds to an *FDR* threshold at the 5% level. (**e**) The genetic correlations between BAG and the 13 complex traits. The red dashed line corresponds to genetic correlation of zero, i.e., the genetic effects on the complex traits are independent of the BAG. AD, Alzheimer’s disease; ASD, Autism spectrum disorder; BIP, Bipolar disorder; BMI: Body mass index; CARD, Coronary artery disease; EDU, education; EPI: Epilepsy; MDD, Major depressive disorder; MS, Multiple sclerosis; PD, Parkinson’s disease; SCZ, Schizophrenia; T2D, Type 2 diabetes.

Several lead SNPs have been reported to be associated with brain structural features and blood cell morphology[32–34]. For example, rs13132853 and rs2819867 have been found as associated with radial diffusivity of white matter microstructure[33] and cortical thickness[32, 34], respectively. In addition, we evaluated the genome-wide genetic heritability 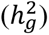 of BAG, and found that the genetic heritability of BAG from our GWAS (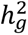 = 0.21, SE = 0.02) was comparable to the results from recent studies (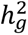 = 0.18, SE = 0.02 from Kaufmann et al.^13^; 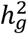 = 0.27, SE = 0.04 from Leonardsen et al.[28]). We also confirmed that our BAG GWAS summary statistics showed high genetic correlation with the results from existing studies (*r_g_* = 0.56, *P* = 2.07e-20 for Kaufmann et al.[13]; *r_g_* = 0.82, *P* = 1.53e-59 for Leonardsen et al.[28], Student’s *t* test).

### Heritability of BAG enriched in regulatory regions and implicated in glial cells

We performed partitioned LD score regression (LDSC)[35] to estimate the enrichment of genetic heritability of BAG in the putative functional genomic regions, as well as with specific brain cell and tissue types (Methods). As expected, we observed significant heritability enrichment in four genomic regions (Figure S7), with the strongest enrichment in super-enhancer regions (enrichment = 2.61, false discovery rate (*FDR)* = 2.97e-08, Student’s *t* test), and enhancer-associated histone modification marker H3K27ac regions (enrichment = 1.87, *FDR* = 5.79e-05, Student’s *t* test). These results suggest that the genome-wide genetic heritability of BAG is particularly enriched in the enhancer regions in human genome.

We then analyzed the enriched genetic signal over genome-wide variants from the BAG GWAS summary statistics dataset within the candidate *cis*-regulatory elements (cCREs) from different brain cell and tissue types using partitioned LDSC[35] (Methods). Given a broad range of neural samples from human brain available in FANTOM5 project[36], we leveraged the annotated active cCREs (i.e., promoters and enhancers) in 12 brain regions from early postnatal and adult periods in this analysis. We found that BAG showed significant heritability enrichment in the active cCREs from diverse brain regions, especially in the adult brain (**Figure 2b**). Additionally, we investigated the heritability enrichment in the cCREs across 43 brain cell types (Table S6)[37]. We found that BAG exhibited stronger heritability enrichment in glial cells than in neurons, which was consistent with previous studies showing that glial cells were more sensitive to brain aging and implicated in degenerative disorders such as AD and Parkinson’s disease (PD)[38–40] (**Figure 2c**).

### BAG is genetically associated with psychiatric and aging-related disorders

Previous studies have indicated that BAG shows variable correlation patterns across diverse brain disorders and traits[13]. However, the susceptibility and causal associations between BAG and brain disorders have remained unclear. The polygenic score (PGS) could be useful in estimating an individual’s genetic predisposition to a specific trait[41]. For each individual in the elder cohort from UKB, we estimated the PGS for each of the 13 complex traits, including neurological disorders, psychiatric disorders, cognitive traits, aging-related disorders, and other background traits (Table S7). We then examined the association between BAG and the PGS across the cohort based on a regression model[42] (Methods). Our findings revealed a significant positive association between BAG and neurological and aging-related disorders, such as AD, PD, and Type 2 diabetes (*FDR* < 0.05, Student’s *t* test; **Figure 2d**), which is consistent with previous studies showing that these disorders could expedite the process of brain aging[43]. Furthermore, BAG was found to be associated with cognitive traits (e.g., education) and psychiatric disorders (e.g., major depressive disorder (MDD) and schizophrenia (SCZ)). These findings were consistent with the observations from the ENIGMA studies showing elevated BAG in patients diagnosed with MDD and SCZ[44, 45]. In addition, BAG displayed weak correlations with height, which was used as the background trait here and assumed not to be implicated in brain aging.

Following the observation of a robust association between BAG and several aging-related traits, we next explored the shared genetic architecture between BAG and these 13 complex traits. We found that BAG showed positive genetic correlation with many brain- and aging-related disorders, as well as cognitive phenotypes (**Figure 2e**). Notably, the genetic correlations between BAG and most of the brain-associated complex traits were generally weak (*r_g_* < 0.1) without reaching statistical significance, which was consistent with previous study[13].

In addition, we examined the underlying causal genetic associations between BAG and the complex traits using Mendelian Randomization (MR)[46] (Methods, Table S8 and TableS9). We identified significant causal genetic relationships (*FDR* < 0.05, *F* test) of two psychiatric and two aging-related disorders on BAG (**Figure 3**, Figure S8, FigureS9, Table S10, and Table S11). For example, the genetic causal effects of schizophrenia on BAG indicate that schizophrenia may cause pathogenic changes of the brain and further lead to accelerated brain aging, which has been supported by previous studies[10]. These results suggest that the accelerated BAG may be the resultant instead of the cause of psychiatric and aging-related disorders.

**Figure 3.**
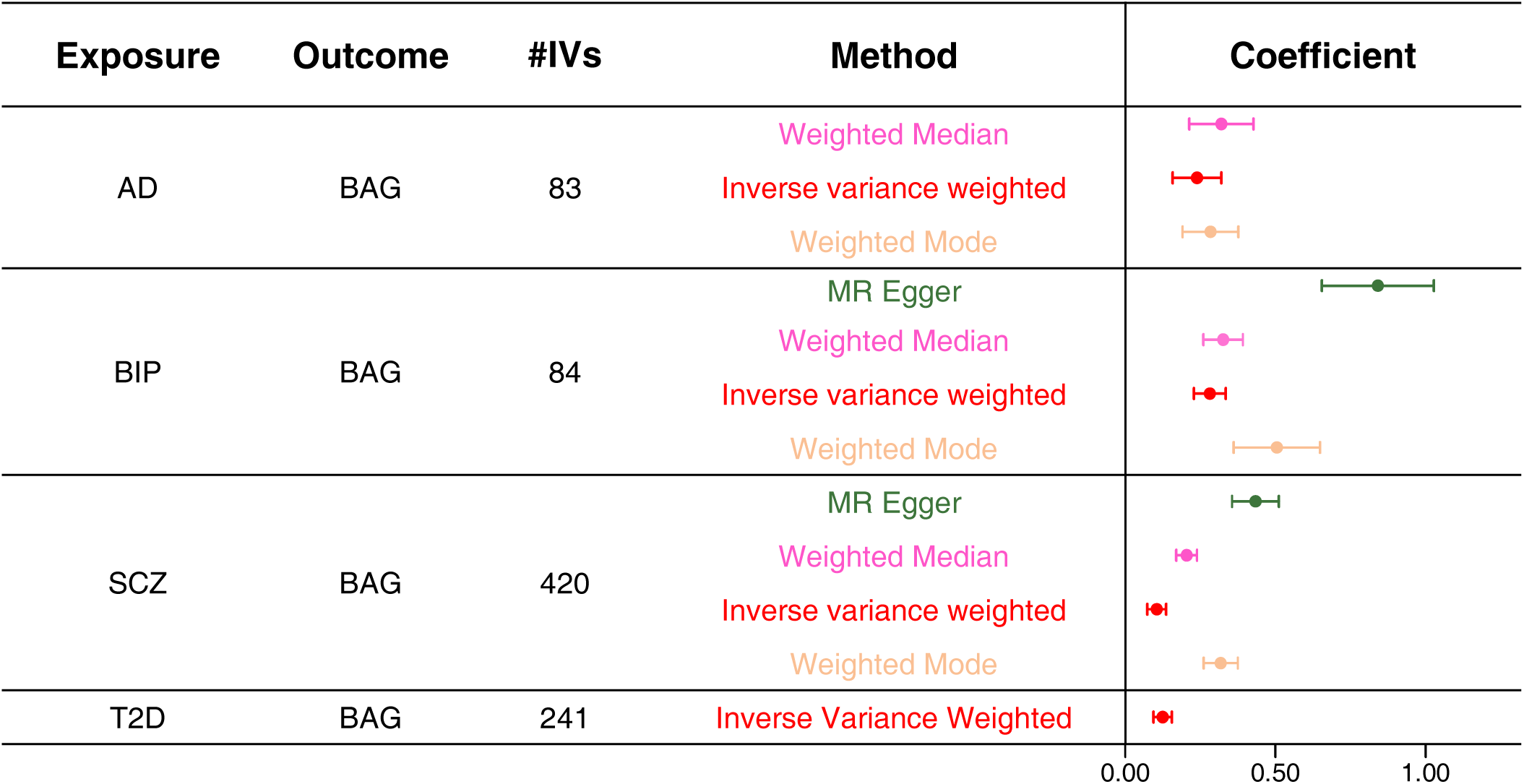
Genetic causal effects of complex traits on BAG. We showed significant causal effects of the complex traits (exposure) on BAG (outcome) after adjusting for multiple testing using the Benjamini-Hochberg procedure to control the *FDR* at the 5% level. #IVs, the number of genetic variants used as instrumental variables. Different MR methods and their regression coefficients are labeled with different colors. AD, Alzheimer’s disease; BIP, Bipolar disorder; SCZ, Schizophrenia; T2D, Type 2 diabetes.

### BAG-associated genes are enriched with aging-related biological pathways

In order to identify genes associated with BAG, we performed gene-based association analysis based on our BAG GWAS summary statistics data using MAGMA[47]. In this step, the genetic heritability of BAG was aggregated to the target genes by proximity, where we identified 48 protein-coding genes (*P* < 2.64e-06, *F* test). To validate the reliability of the MAGMA-derived gene set, we also mapped the significant genetic variants (*P* < 5e-08 in the BAG GWAS summary statistics) to their target genes based on eQTL and chromatin interaction (Hi-C) data from human brain using FUMA[14], resulting in the identification of an additional set of 64 and 78 genes, respectively[48]. The gene sets prioritized using functional genomic data showed significant (Figure S10, *P* < 2.2e-16 for eQTL and Hi-C, Fisher’s exact test) overlap with the MAGMA-derived gene set (Table S12). Some well-known BAG-associated genes *RUNX2, MAPT, ANKRD11*, and *KLF3* were identified by both approaches[13, 14, 28].

We then performed a gene-set enrichment analysis to identify the enriched biological functions of these BAG-associated genes. We found that the BAG-associated genes were enriched with brain aging-related biological pathways (**Figure 4a**), such as Wnt signaling pathway, which was critical for synaptic plasticity and maintenance in the adult brain[49]. Dysregulation of Wnt signaling pathway has been implicated in the functional decline during aging and pathogenesis of neurodegenerative diseases such as AD[50]. As expected, the BAG-associated genes are predominantly enriched with biological functions such as bacterial lipoprotein and synaptic transmission (**Figure 4a**). For example, bacterial lipoprotein has been identified as a key factor implicated in neuroinflammation and neurodegenerative diseases[51–53]. These enriched biological functions suggest that the BAG-associated genes we identified are biologically meaningful, and there is a substantial association between these genes and the process of brain aging.

**Figure 4.**
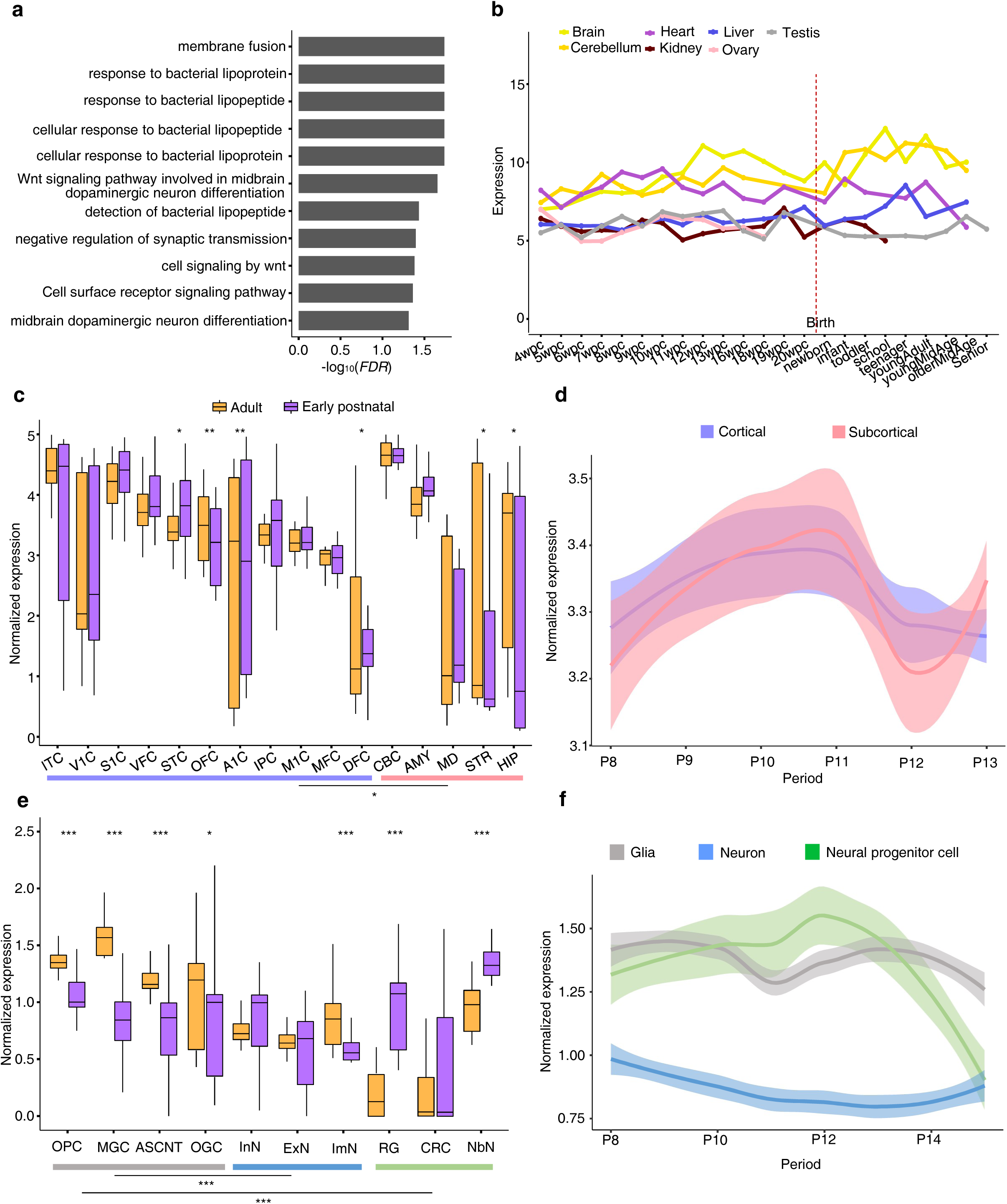
Enriched biological functions and expression dynamic patterns of BAG-associated genes. (**a**) Significantly enriched biological functions of BAG-associated genes (FDR < 0.05). (**b**) Expression dynamics of BAG-associated genes in different tissue across lifespan. The expression data of BAG-associated genes were obtained from Cardoso-Moreira *et al*.[54] (**c**) Expression of BAG-associated genes in adult versus early postnatal stage in different brain regions. *, *P* < 0.05; ***, *P* < 0.001; Wilcoxon test. The 16 different brain regions were separated as cortical (OFC, orbital prefrontal cortex; STC, superior temporal cortex; VFC, ventrolateral prefrontal cortex; MFC, medial prefrontal cortex; IPC, inferior posterior parietal cortex; DFC, dorsolateral prefrontal cortex; S1C, primary somatosensory cortex; V1C, primary visual cortex; ITC, inferior temporal cortex; M1C, primary motor cortex; A1C, primary auditory cortex) and subcortical (AMY, amygdala; STR, striatum; MD, mediodorsal nucleus of thalamus; CBC, cerebellar cortex; HIP, hippocampus) ones. (**d**) Expression dynamics of BAG-associated genes in cortical and subcortical regions. The expression levels of BAG-associated genes were obtained from Zhu *et.al.*[55] (**e**) Expression of BAG-associated genes in glial (grey) versus neuronal (blue) and neural progenitor (green) cell types. Glial cell types: OGC, oligodendrocytes; MGC, microglia; ASCNT, astrocytes; OPC, oligodendrocyte progenitor cells. Neuronal cell types: InN, inhibitory neurons; ExN, excitatory neurons; ImN, immature neurons. Neural progenitor cell: RG, radial glia; CRC, cajal-retzius cells; NbN, newborns neuron. PCW, postconceptional weeks. (**f**) Expression dynamics of BAG-associated genes in glial, neuronal and neural progenitor cell types. The expression levels of BAG-associated genes were obtained from our STAB2 database[58]. The LOESS plots show smooth curves with 95% confidence bands. P8, Birth ≤ Age < 6 Months; P9, 6 Months ≤ Age < 1 Year; P10, 1 ≤ Age < 6 Years; P11, 6 ≤ Age < 12 Years; P12, 12 ≤ Age < 20 Years; P13, 20 ≤ Age < 40 Years; P14, 40 ≤ Age < 60 Years.

### BAG-associated genes show elevated expression in adult brain regions and glial cells

Hypothesizing that the BAG-associated genes are functional in regulating aging in brain and/or whole body, we investigated the expression dynamics of the BAG-associated genes using transcriptomic datasets from seven different tissue types (including brain, cerebellum and another five non-brain tissue types) across human lifespan[54]. The BAG-associated genes exhibited consistently higher expression levels in the cerebrum and cerebellum compared to non-brain tissue types throughout the majority of the lifespan (**Figure 4b**). Although BAG can represent the process of aging in the whole body, these results suggest that the BAG-associated genes are relatively more associated with and may be functional in human brain.

Next, we systematically compared the expression levels of BAG-associated genes of early postnatal and adult periods in 16 different brain regions[55]. We found that BAG-associated genes showed significantly higher expression in the adult than in the early postnatal period in four brain regions, including primary auditory cortex, orbital prefrontal cortex, striatum, and hippocampus (*P* < 0.05, Wilcoxon test; **Figure 4c**), that are related to the functions of perception and memory[56, 57]. We also noticed that BAG-associated genes generally have significantly higher expression in cortical regions than in subcortical regions during aging (*P* = 0.03, Wilcoxon test; **Figure 4c**). Given this observation, we then systematically characterized the expression dynamics of BAG-associated genes in cortical and subcortical regions from birth to adulthood. The cortical regions showed an earlier expression elevation of BAG-associated genes than the subcortical regions from the beginning of adulthood (**Figure 4d**), suggesting that the cortical regions may age earlier than the subcortical regions[2].

We further explored the expression patterns of BAG-associated genes in brain cell types, utilizing the single-cell RNA-seq data from our STAB2 database[58]. Consistent with our previous results from partitioned heritability enrichment in brain cell types, BAG-associated genes were significantly highly expressed in adult brain cells (*P* < 2.2e-16, Wilcoxon test), particularly in glial cells such as oligodendrocytes and microglia (**Figure 4e**). In addition, during aging, BAG-associated genes showed consistently higher expression levels in glia than in neurons (**Figure 4f**). These observations confirmed that our previous results that glial cells were more sensitive to the process of aging.

### BAG-associated genes exhibit strong functional correlations with aging-related epigenetic clock

When predicting brain age, a crucial aspect is accommodating the association of BAG with the biological aging processes. However, there have been limited studies aiming to explicate their connections by utilizing BAG as a marker and assessing its correlation with cognitive phenotypes[59]. Here, we established a link between brain aging and biological aging through the regulatory mechanism of epigenetic clock, defined as a specific collection of CpG sites whose DNA methylation levels can be used to estimate a subject’s age[60]. The methylation age gap (MAG), defined as the gap between the predicted biological age based on methylation signal in blood and the chronological age, has been widely used as a biomarker of aging processes[61]. Briefly, we applied a predefined epigenetic clock model and our ACN model to predict the methylation age and brain age respectively based on the ADNI cohort[62, 63], and then calculated the BAG and MAG for each individual (Methods). The BAG and MAG showed a concordant pattern in patients diagnosed with AD, which had a significant positive BAG compared to healthy participants (Figure S11). Upon investigating the correlation between the BAG and the MAG, we observed a significant association between BAG and MAG (correlation coefficient *r* = 0.56, *P* < 2.2e-16, Pearson correlation test; **Figure 5a**), indicating that our predicted brain age was reliable to represent the chronological age and can be used to characterize the potential aging processes. Furthermore, we identified a set of MAG-associated genes based on the epigenetic clock, followed by evaluating the similarity between BAG-and MAG-associated genes by computing their functional similarity (Methods). We observed a high degree of functional similarity between these two gene sets (GOSim[64] = 0.80).

**Figure 5.**
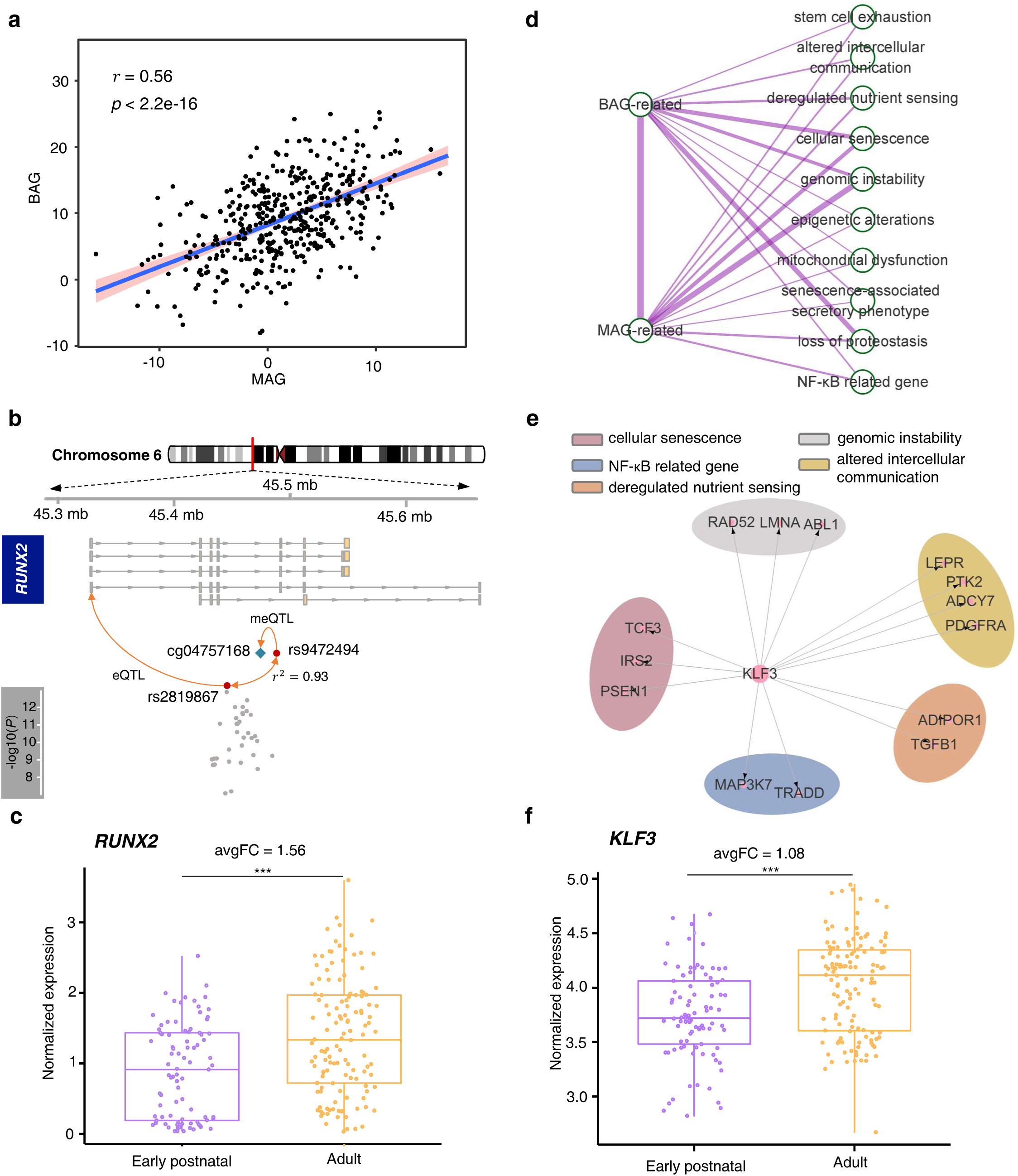
Genetic association between BAG and MAG. (**a**) The association between BAG and methylation age gap (MAG) in ADNI dataset. (**b**) Shared genetic loci between BAG and the epigenetic clock, located in *RUNX2*. (**c**) Expression of *RUNX2* in brain regions during early postnatal versus adult stage. ***, *P* < 0.001. (**d**) The overlap between BAG-/MAG-associated genes and different aging-related biological pathways (Methods). The width of the lines indicates the correlation strength among different gene sets. (**e**) An example showing regulatory target genes of *KLF3* in different aging-related biological pathways. (**f**) Expression of *KLF3* in brain regions during early postnatal versus adult stage. ***, *P* < 0.001. FC, fold change of gene expression between adult and early postnatal period.

Here, we exemplified *RUNX2*, which has been identified as both a BAG-associated and MAG-associated gene in our analysis, to show the complex regulatory relationships between BAG and the epigenetic clock (**Figure 5b**). *RUNX2* has been identified as the regulatory target of the eQTL SNP rs2819867[65], which is also identified as the BAG lead SNP in our study, and in strong LD with a nearby SNP rs9472494 (*r*^2^ = 0.93). Interestingly, the SNP rs9472494 has been identified as a DNA methylation QTL of cg04757168[66], which is located in the gene *RUNX2*. RUNX2 has been reported to be implicated in the pathogenesis of aging-related disorders, such as PD[67]. Consistent with these results, we found that *RUNX2* showed elevated expression in the adult brain than in the early postnatal brain (fold change (FC) = 1.56, *P* = 7.6e-06, Wilcoxon test; **Figure 5c**).

As we have discovered that BAG-associated genes showed significantly functional overlap with MAG-associated genes and could be linked to aging processes, we raised the question of which aging-related biological pathways were implicated in both BAG- and MAG-associated genes. To this end, we first collected aging-related gene sets, which were organized into 10 different categories of aging-related biological functions or pathways[68]. We then examined the potential regulatory relationships between BAG-associated genes, MAG-associated genes, and aging-related biological pathways separately in the transcriptional regulatory networks (TRNs)[69]. BAG-associated genes showed higher connectivity with MAG-associated genes in the TRNs compared to other genes (mean connectivity score (*C_s_*) = 1.64, Figure S12; Methods). Both BAG- and MAG-associated genes were highly implicated in the aging-related biological functions or pathways, particularly in genomic instability and cellular senescence (**Figure 5d**). Intriguingly, *KLF3*, a key TF in the BAG-associated genes, was found as the regulator of numerous aging-related genes, particularly those involved in intercellular communication (e.g., *LEPR*, *PTK2*, *ADCY7*) and cell senescence (e.g., *IRS2*, *PSEN1*) (**Figure 5e**), which have been implicated in synapse formation, lipid metabolism, and insulin resistance[70, 71]. Additionally, *KLF3* showed significantly higher expression in the adult brain (FC = 1.08, *P* = 3.4e-06, Wilcoxon test; **Figure 5f**), and regulated 23 MAG-associated genes, which were involved in regulating immune-related functions and cell development[72–75].

Taken together, these findings suggest that the aging processes estimated by the epigenetic signal in blood share certain biological functions and/or pathways with the aging in brain, particularly for the functional categories of immune senescence and cell senescence-associated pathways.

### Genetic pleiotropy of BAG-associated genes in brain disorders

The aging trajectory of the brain has been associated with brain disorders in previous research[2]. Nonetheless, the functional mechanisms bridging brain aging and brain disorders remain to be comprehensively understood. We thus investigated the potential biological association between brain aging and neurological disorders, utilizing molecular network analysis to capture the gene associations between BAG-associated genes and seven different brain disorders[76] (Table S13; Methods).

We considered three different types of molecular networks in the human brain to uncover the potential association with brain disorders, including TRNs, protein-protein interaction (PPI) networks, and gene co-expression networks (Methods). Our results highlighted that BAG-associated genes and many risk genes associated with neurological disorders were more strongly enriched in the TRNs compared to the gene co-expression networks and PPI networks (**Figure 6a**, Figure S13, Figure S14, and Figure S15). These results suggest that the TRNs in human brain may better represent the potential regulatory relationships concerning brain aging and disorders[77].

**Figure 6.**
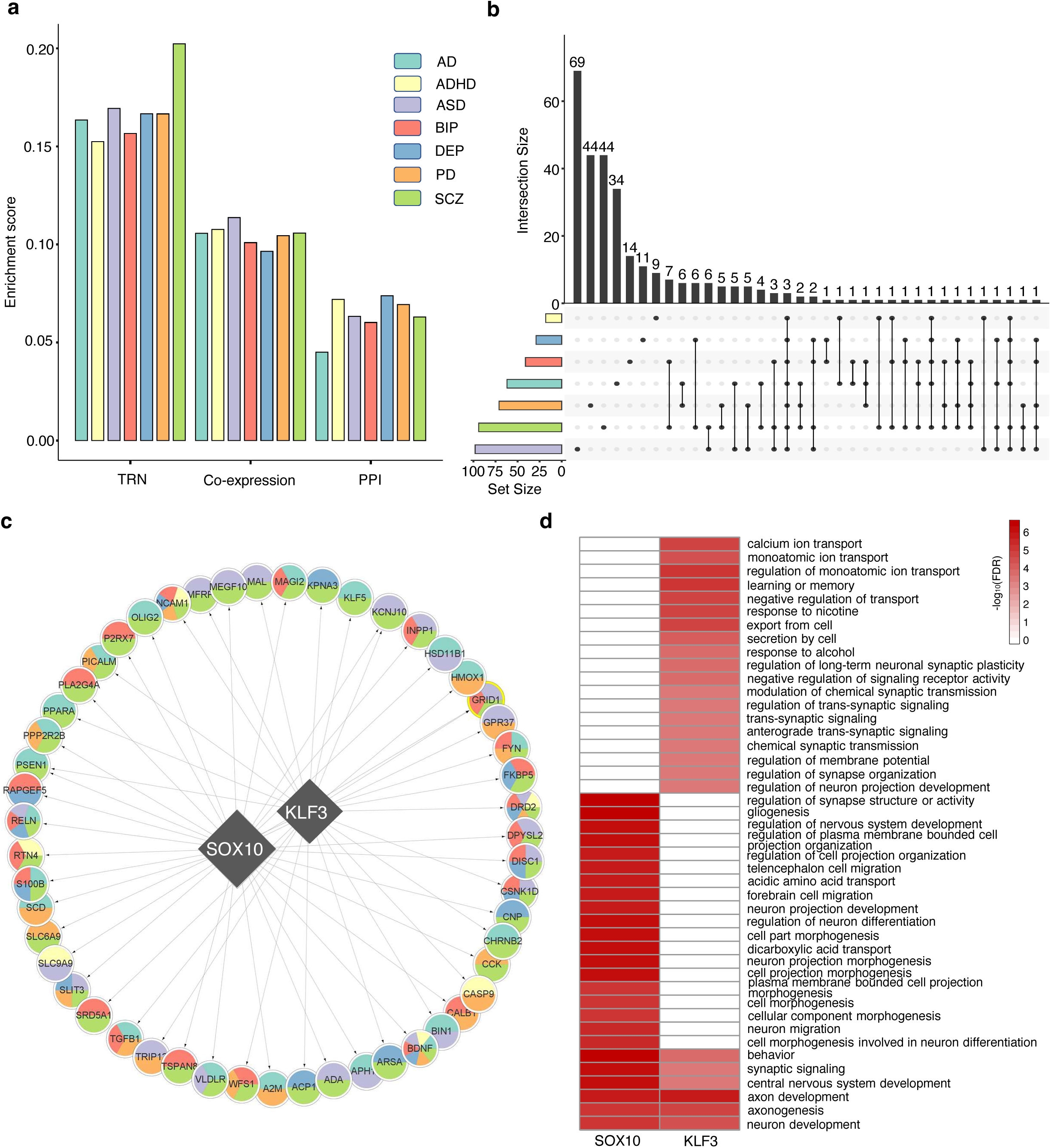
The pleiotropy between BAG-associated genes and risk genes of brain disorders in molecular networks. (**a**) Enrichment of BAG-associated genes with risk genes of diverse brain disorders in molecular networks. The enrichment is estimated using the Jaccard distance index, for which the higher value indicates greater similarity. The enrichment analysis was performed based on different networks, including TRNs, PPI networks, and gene co-expression networks from the human brain (Methods). AD, Alzheimer’s disease; ADHD, Attention deficit hyperactivity disorder; ASD, Autism spectrum disorder; BIP, Bipolar disorder; MDD, Major depressive disorder; PD, Parkinson’s disease; SCZ, Schizophrenia. (**b**) The overlap between the regulatory target genes of BAG-associated genes in the brain TRNs for diverse brain disorders. (**c**) Delineation of pivotal hub modules for BAG-associated genes, where diamonds symbolize BAG-associated genes, circles denote risk genes of brain disorders, and the varied colors in the pie chart represent the genes that are common across different brain disorders. (**d**) Significantly enriched biological functions of the target genes regulated by the BAG-associated hub TFs, *KLF3* and *SOX10*.

To explore the impact of brain aging on diverse brain disorders, we first examined the pleiotropy of disorder-associated risk genes modulated by BAG-associated genes within the brain TRNs. Our results indicated that 22.68% of the risk genes that had interactions with BAG-associated genes were involved in at least two brain disorders (**Figure 6b**). Intriguingly, we detected two major hub modules (ranked by degree) regulated by *KLF3* and *SOX10*, where their regulatory target genes were enriched with risk genes of diverse brain disorders (**Figure 6c**). The risk genes regulated by *KLF3* and *SOX10* were significantly enriched with the biological functions of brain development (e.g., central nervous system development and neuron development) and cognition (e.g., learning or memory) (**Figure 6d**).

We also explored the interactions between BAG-associated genes and risk genes associated with diverse disorders within the co-expression and PPI networks in human brain(Table S14). Consistent with the results from the brain TRNs, 34.10% and 50.0% of the risk genes that had interactions with BAG-associated genes were associated with at least two brain disorders in the co-expression and PPI networks from human brain, respectively (Figure S16 and Figure S17).

Given that our previous MR results have revealed causal effects of psychiatric disorders on BAG, we also examined the regulatory influence of risk genes on BAG-associated genes. As expected, we found that the BAG-associated gene *MAPT* was a regulatory target of several risk genes associated with psychiatric disorders, such as *CLOCK* and *ARNTL*[78]. *MAPT* has been reported as functional in regulating cognition[79]. These results suggest that certain psychiatric conditions may accelerate brain aging by affecting the underlying mechanisms of cognition[80, 81].

## Discussion

Our study reports the catalogue of BAG-associated genetic variants and genes, which were identified by the ACN model, a novel computational framework for brain age prediction. With this catalogue, we interrogate the expression patterns and regulatory mechanisms of the BAG-associated genes and uncover their associations with epigenetic clock and aging-related biological pathways.

Previous studies suggest that the aging process of the brain exhibits gender differences, in which female brains are generally younger than male ones[82, 83]. Although gender has been well considered as a confounding factor when predicting brain age in most established computational models, it should also be noted that there exist indeed sex-biased differences in the brain structure[30], which is usually ignored in most current models for brain age prediction. To overcome this difficulty, we proposed ACN, an adversarial model to minimize the influence of sex-biased differences of brain structure in brain age prediction. The main advantage of our ACN model is that we utilized a domain-adversarial training strategy to reduce the influence of the noise and gender effects from different data sites, and make the stratified features more relevant to the real brain age (Figure S18). We found that this approach could improve the accuracy in brain age prediction compared to alternative models, which laid a solid foundation for the downstream analysis of GWAS and prioritization of BAG-associated genes.

We estimated the genetic correlations and causal relationshipes between BAG and different brain-/aging-related disorders. As expected, BAG exhibited complex genetic associations with diverse brain-/aging-related disorders instead of the background tratis such as height, suggesting that the BAG estimation by our ACN model was biologically meaningful and could reflect brain- and age-related physiological changes. Notably, our results suggest that psychiatric (SCZ and BIP) and aging-related (AD and T2D) disorders could induce accelerated brain aging. We identified nine novel BAG-associated genetic variants. Furthermore, we prioritized 44 novel genes that were associated to BAG and showed that these BAG-associated genes have significantly elevated expression in brain aging-related regions and cell types (e.g., adult hippocampus and glial cells). Some BAG-associated genes (e.g., *RUNX2*) showed genetic overlap with the epigenetic clock and aging-related biological pathways (e.g., cell senescence). Finally, we examined the potential regulatory relationships between BAG-associated genes and risk genes of brain disorders in molecular networks. We highlighted two pleiotropic BAG-associated TFs (*KLF3* and *SOX10*) that potentially play key regulatory roles in different brain disorders.

Brain aging is a complex biological process, and BAG, derived from the MRI estimations, can be used as a reliable measure for quantitatively assessing the process of brain aging. However, BAG is usually predicted based on computational models using different brain imaging data, and its relevance to the biological processes of aging remains vague. Great efforts have been made to determine whether the predicted BAG is physiologically reliable by examining its relations with brain-associated phenotypes, such as cognitive ability or mental disorders. Over the past decade, epigenetic clock has emerged as a highly accurate molecular indicator of biological age in humans[84]. In this study, we proposed to validate BAG as a novel indicator of brain aging by revealing the putative connection between BAG and biological aging through the epigenetic clock. Our study showed that BAG-associated genes have strong correlations and functional overlaps with the genes involved in epigenetic clock.

We revealed that the genetic heritability of BAG was significantly enriched in functional regulatory regions and glial cells. Glial cells play critical roles in regulating the molecular processes including neuroinflammation and synaptic pruning. Dysregulation of these processes has been implicated in neurodegenerative diseases and aging-associated cognitive decline[85–87]. Our results suggest that accelerated brain aging has genetic association with certain psychiatric and aging-associated disorders, such as AD and schizophrenia, both of which have been revealed as linked to the dysfunction of glial cells[88, 89]. Taken together, these findings indicate that glial cells may play key roles in regulating brain aging, and the dysfunction of glial cells could induce diverse brain- and aging-associated disorders.

Previous studies have revealed that brain disorders can expedite brain aging and uncovered significant independent loci associated with BAG (e.g., rs2106786 and rs2790102)[13, 28]. However, the associations between BAG, brain aging and brain disorders at gene level have not been well characterized. In our study, we validated well-known genes and identified some novel genes that were associated with BAG. We further examined the enrichment of BAG-associated genes with the risk genes of diverse brain disorders in biological networks, and found two common gene modules (*KLF3*, *SOX10*). *KLF3* has been associated with various aging-related biological pathways and can regulate numerous risk genes of diverse brain disorders, indicating its strong pleiotropy in brain disorders. The regulatory target genes of *KLF3* (e.g., *TGF-β1*, *FYN,* and *DRD2*) have been shown to participate in the regulatory processes of neurodevelopment and inflammation[90–94]. *SOX10* can significantly interacts with some risk genes associated with neuroinflammation implicated in neurodegenerative disorders, such as *RNF11* and *LAMP1*[95, 96] (Figure S17). Interestingly, we also found that BAG-associated genes *KLF3*, *MAPT*, *WNT3* and *RUNX2* were all associated with the Wnt signaling pathway, which played critical roles in the regulation of brain development, the aging process, and a variety of neurological disorders[97, 98]. In conclusion, our results found that multiple disease risk genes that interact with BAG-associated genes are associated with Wnt signaling pathway and neuroinflammation, while BAG-associated variants as well as genes are significantly enriched in neuroinflammation related brain cells, glial cells. These results suggest that the neuroinflammation and Wnt signaling pathway are implicated in abnormal aging process of human brain and may be involved in the pathogenesis of psychiatric and neurodegenerative disorders[99–101].

The BAG metric used in this study was developed from structural MRI data collected from a large cross-sectional dataset of individuals spanning a wide age range, and it encapsulates a prediction error from a deep learning model. Therefore, its limitations include both noise (i.e., model accuracy deficiencies, and inadequate data quality) and physiology (i.e., deviations from standard aging trajectories). Despite considering site effects and healthy conditions, care is necessary when estimating the aging trajectory of different cohorts with a limited population size. Given that the genetic architecture of BAG could be multifarious and population-specific, we only used data from individuals with European ancestry. Future studies may need to be repeated in other races as well as in larger cohorts. Furthermore, while evidence has been provided that BAG can serve as a biomarker to delineate the brain aging process, it may demonstrate a stronger correlation with structural changes in the brain. In future work, the aging trajectory of the brain can be depicted based on different brain function modalities and in combination with multi-omics data of the brain.

In conclusion, we investigated the genetic architecture of BAG, followed by prioritizing the BAG-associated genes and revealing their biological significance and relations to brain disorders at functional genomic levels. Our findings suggest the BAG could be used as an effective biomarker to characterize the process of brain aging and brain disorders.

## Methods

### Collection and preprocessing of structural MR images

We compiled a dataset of T1-weighted MR images from five independent data sites (Table S1): Southwest University Adult Lifespan Dataset (SALD)[21], Citigroup Biomedical Imaging Center (CBIC)[22], Australian Imaging Biomarkers and Lifestyle Study of Ageing (AIBL)[23], IXI (https://brain-development.org/ixi-dataset/), and Open Access Series of Imaging Studies (OASIS)[24]. In total, this compiled dataset included 2,011 healthy individuals (ages ranging from 5 to 94 years), and was used for the model training and testing. During model training and testing, the MR images were split into 70% (N=1,407) for training, 10% (N=201) for validation, and 20% (N=403) for testing.

We also leveraged another two MR image datasets for model application: the UK Biobank (UKB) dataset (comprising 38,702 individuals with ages ranging from 45 to 85 years), and the Alzheimer’s Disease Neuroimaging Initiative (ADNI) dataset (comprising 437 individuals with ages ranging from 55 to 91 years). The UKB dataset and ADNI dataset were applied on the GWAS analysis of BAG and the association analysis between BAG and epigenetic clock of aging, respectively.

We performed a four-step procedure for the preprocessing of the raw MR image data: 1) the N4 bias correction was applied[102]; (2) skull strip, which involved skull removal and brain extraction using FSL; (3) nonlinear registration of the image to the standard MNI space using ANTs[103]; (4) normalization of voxel value, with all voxel sizes aligned to 1.5×1.5×1.5 mm^3^; (5) segmentation of the image into grey matter, white matter and cerebral spinal fluid. Following preprocessing, the voxel resolution was 121 × 145 × 121.

### Development of ACN model for brain age prediction

We developed ACN model, a novel computational model, to predict brain age. This model comprises an encoder, a task classification (regression) module, and mixed modules. The encoder selectively extracts age-related features while protecting the feature domain from confounding variables such as data site and gender information. Details of brain age prediction are shown in Figure S1.

In this study, we proposed a model that can integrate irrelevant factor mixed blocks by harnessing the principles of adversarial learning. These mixed factor tasks hold a negative correlation with the principal task of brain age prediction. To achieve the brain age prediction (Baseline model), two approaches were utilized. Initially, we used conventional regression with L1 loss to constrain the output. However, due to the heterogeneous distribution of age, a distribution constraint was also included. For each age label *y*, the cumulative distribution function of a normal distribution was employed to convert this into a probability distribution, resulting in a 90-dimensional vector (5-94). The conversion function is illustrated below:

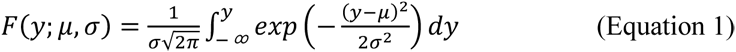

where the *μ* and *σ*^2^ were the expectation and variance for the age labels.

The output layer comprises 90 numbers that represent the predicted probability that the subject’s age falls into a one-year age interval between 5 and 94. The KL divergence computes the correlation for the probability distribution of the label and prediction as the loss. Consequently, the loss of brain age prediction can be formulated as follows:

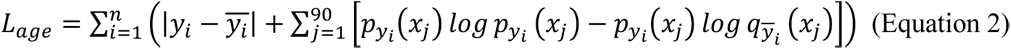

where *y* is the true label, and *ȳ* denotes the predicted value of the model. With certifying the effectiveness of the two loss functions, we have compared the result of model with different loss constrain (Table S2).

In contrast, the mixed module might incorporate tasks that potentially affect the generalization of the main task, such as gender and different datasets. To hinder the encoder from extracting features associated with these factors and to align the domains based on gender and data site, we extended the domain adaptation method to the mixed module. Specifically, a gradient inversion layer was placed ahead of the mixed block to constrain the feature encoder. The gradient update method mirrored that of DANN[20]. In this case, the loss for confusion factors was cross-entropy, which can be expressed as follows:

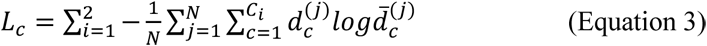

where *i* was the number of different factors, *C_i_* denoted dimension for different classification model output, *d̄* denoted the output of the classification model. The learning process minimizes the loss function that is written as follows:

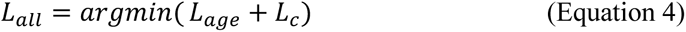

To evaluate the effectiveness of the model featuring a mixed factor module of gender and data site, we compared its results with those of a model without this module (**Table 1**).

The loss function was optimized by Adam[104] with a learning rate of 1e-04 and a batch size of 12. The training process consisted of 200 epochs, with each epoch including a training step and a validation step. Our model implementation utilized approximately 20 GB of memory. The training time, employing an AMD EPYC 7763 64-Core processor with 2T of RAM and an NVIDIA Tesla A100 80GB GPU, took approximately 8 hours.

### Applying the pre-trained ACN model to the UKB cohort

We adapted our pre-trained ACN model to the elder cohort from the UKB dataset through transfer learning. We first selected approximately 10% (N = 3,000) of the individuals from the UKB cohort for model fine-tuning. These individuals were divided into three subsets: 70% for training, 10% for validation, and 20% for testing. The model was trained on the selected individuals (N = 3,000) and fine-tuned over 100 epochs. The fine-tuned model showed improved performance on the test set, achieving a MAE of 2.51 and a PCC of 0.91. We then applied this optimized ACN model to the remaining individuals (N = 35,702) from the UKB cohort to predict their brain ages, followed by computing their BAG scores.

### GWAS summary statistics data of complex traits

We collated a set of GWAS summary statistics for 13 complex traits (Table S7). Most of these GWAS summary statistics datasets were recently published and available, as well as based on meta-analyses using extensive sample sizes of European ancestry.

The sample size for each of the GWAS summary statistics datasets was shown as follows:

- Neurological disorders: Alzheimer’s disease (AD)[105], 7,428 cases and 429,961 controls; Epilepsy (EPI)[106], 15,212 cases and 29,677 controls; Multiple sclerosis (MS)[107], 7,428 cases and 429,961 controls; Parkinson’s disease (PD)[108], 33,674 cases and 449,056 controls.
- Psychiatric disorders: Autism spectrum disorder (ASD)[109], 18,381 cases and 27,969 controls; Bipolar disorder (BIP)[110], 41,917 cases and 371,549 controls; Major depressive disorder (MDD)[111], 135,458 cases and 344,901 controls; Schizophrenia (SCZ)[112], 76,755 cases and 243,649 controls.
- Cognitive traits: Education (EDU)[113], 766345 individuals.
- Other aging-related disorders: Coronary artery disease (CARD)[114], 22,233 cases and 64,762 controls; Type 2 diabetes (T2D)[115], 22,233 cases and 64,762 controls.
- Background traits: Body Mass Index (BMI)[116], 694,649 individuals; Height[117], 450,000 individuals;

### Genetic data preprocessing and association analysis

Genetic analyses were performed on individuals from the UKB with European ancestry, who also had genotypes and T1-weight MRI image data. Standard quality control procedures were then applied to the UKB v3 imputed genetic data[118]. These procedures included the following steps: (1) exclusion of individuals with failed genotyping, abnormal heterozygosity status, or withdrawn consents. (2) removal of participants genetically related—up to the third degree—to another participant, as inferred by kinship coefficients implemented in PLINK[119]; (3) elimination of variants with a minor allele frequency below 0.01; (4) removal of variants with a genotype missing rate exceeding 10%; (5) exclusion of variants failing the Hardy-Weinberg equilibrium test at the 1e-07 level; 6) elimination of variants with an imputation INFO score below 0.8. Post quality control, we retained 38,702 individuals and 9,055,103 variants.

In terms of the genome-wide association analysis (GWAS), we performed the BAG GWAS analysis using GCTA, based on the generalized linear mixed mode[120], with adjustments for some covariates. This included age (at imaging), age squared, sex, sites, age-sex interaction, age-squared-sex interaction, intracranial volume, imaging center, as well as the top 20 genetic principal components provided by UKB (Data-Field 22009). The *P*-value threshold for selecting the significant GWAS tag variants is 5e-08.

We leveraged partitioned LDSC[35] to estimate the heritability enrichment in the regulatory elements active in each brain cell and tissue type. Briefly, LDSC regresses GWAS χ2 statistics on SNPs’ LD scores, reflecting the degree to which each SNP is correlated with its surrounding SNPs[121]. The LD Score regression intercept of BAG GWAS was close to one (intercept = 0.97, standard error (SE) = 6.8e-03), indicating the absence of genomic inflation of test statistics due to confounding factors. Moreover, the pre-calculated genome-wide LD scores were obtained from LDSC (https://data.broadinstitute.org/alkesgroup/LDSCORE/). The LD scores were calculated based on data from individuals of European ancestry from the 1000 Genomes Project[122]. We further removed SNPs which were not annotated in HapMap3[123] or located in the major histocompatibility complex regions.

### Computing polygenic scores and correlation with BAG

A polygenic score (PGS) represents an estimate of an individual’s genetic predisposition for a given trait. For each individual from the UKB cohort, we computed the PGS for each trait based on the genotype data of the individual and the GWAS summary statistics dataset using PRSice-2[124]. Notably, we double-checked the sample metadata of the collected GWAS summary statistics to ensure that there was not sample overlap between the UKB cohort and the GWAS cohorts used in our study (Table S7).

Prior to PGS computation, we performed quality control on the UKB genotype data, e.g., removing the duplicate and ambiguous SNPs, clumping to account for linkage disequilibrium, pruning highly correlated SNPs, and filtering out high-heterozygosity samples. We applied the P-thresholding approach in PRSice-2 for PGS computation, for which the *P*-value thresholds of SNPs were tested ranging from 5e-08 to 1[124]. We then performed linear regression analyses to examine the correlation between PGS and BAG while adjusting several covariates including age, sex, intracranial volume, and the top 10 genetic principal components. The optimal threshold for each trait was chosen by maximizing the incremental R² in the linear regression model. All P-values were converted into FDR by conducting multiple testing correction.

### Mendelian randomization

We first confirmed that there was not sample overlap between the UKB cohort and the GWAS cohorts used in the Mendelian randomization (MR) analysis. We then preprocessed the GWAS summary statistics datasets in accordance with standard MR preprocessing procedures. In the exposure GWAS, the genetic variants were selected based on a genome-wide significance threshold of 5e-08. To ensure the independence of genetic variants included in the subsequent MR analysis, we applied LD-based clumping with an *r*^2^ threshold of 0.1, a window size of 250 kb, and utilized the European ancestry data from the 1000 Genomes Project as the reference panel[122]. The MR procedure was performed using the R package TwoSampleMR (version 0.4.26, https://mrcieu.github.io/TwoSampleMR/).

We also performed sensitivity analysis on the MR results. We used the *harmonise_data* function in the TwoSampleMR to harmonize the effect alleles and SNP effects between the exposures and outcomes. We used the *mr_heterogeneity* function (*P* < 0.05) and the *mr_pleiotropy_test* function (*P* < 0.05) to detect the heterogeneity (Table S13) and horizontal pleiotropy (Table S14) in the MR results, respectively. We also used the *mr_leaveoneout* function to test the sensitivity between the instrumental variables and the MR results.

### Prioritizing BAG-associated genes

In the study of the BAG GWAS, MAGMA (version 1.08)[47] was used to perform a gene-based association analysis for 18,796 protein-coding genes. The default MAGMA parameter settings were applied, with a zero-window size around each gene. Subsequently, FUMA functional annotation and mapping analysis were performed, which involved annotating variants with their biological functionality and linking them to candidate target genes through a combination of eQTL and 3D chromatin interaction mappings. Brain-related tissues/cells were selected for all options, and default parameters were used[48]. We used ToppGene[134] to identify the significantly enriched (FDR < 0.05) Gene Ontology terms of the BAG-associated genes.

### Collection and analysis of methylation data

We procured the DNA methylation data from the ADNI cohort[62], employing the Illumina EPIC chips. Subsequently, we used the chip analysis methylation pipeline (CHAMP) to preprocess the methylation with default parameters[125]. These steps included: (1) filtering out low-quality probes; (2) quality control of samples; (3) signal normalization; (4) removing batch effects and regressing sex. After these steps, the remaining samples had an average CpG call of 695,127. Finally, we selected 437 individuals with brain MR imaging data for biological age prediction.

We applied a recently published elastic network model to predict the biological ages based on the blood methylation data[63]. This model was trained on 13,402 samples with blood methylation data, identifying 514 CpGs as the epigenetic clock. For this study, the epigenetic clock was extracted as the feature, and the pre-trained elastic network model was used to predict the methylation age for each participant. Additionally, the epigenetic clock was mapped to 351 associated genes using the Illumina EPIC array annotation file.

### Collection of risk gene for brain disorders

Risk genes associated with brain disorders were compiled from various resources: (1) risk genes of ADHD were sourced from the ADHDgene database (http://adhd.psych.ac.cn), selecting only those with support from at least 60% of all studies included in the database[126]; (2) risk genes of ASD were downloaded from the AutDB database (http://autism.mindspec.org/autdb) and were supplemented with risk genes from recent studies[127, 128]; (3) risk genes of SCZ were obtained from the SZGene database (http://www.szgene.org/) and from research by Wang *et al.*[65, 129]; (4) risk genes of BIP were gathered from DisGeNet[130]; (5) risk genes of MDD were downloaded from the Polygenic Pathways database (http://www.polygenicpathways.co.uk/depression.htm); (6) risk genes of AD were obtained from the ALzGene database (http://www.alzgene.org)[131]; (7) risk genes of PD were obtained from the PDGene database (http://www.pdgene.org)[132]. The full list of these risk genes can be found in Table S11.

### Collection of molecular networks

Three extensive molecular networks were utilized in this study to investigate the association between BAG and brain disorders: (1) Brain-specific TRNs were obtained from Pearl *et al.*[69], which included 741 transcription factors (TFs) and 11,092 target genes; (2) Brain-active PPI networks were reconstructed by first downloading the global PPI networks from STRING (https://string-db.org), followed by retaining of protein pairs with physical interaction scores over 700, and proteins active in the adult human brain[55] (expression value > 0), which finally included 8,568 proteins and 114,892 interaction edges (Table S12); (3) Gene co-expression networks in adult brain were generated by first removing lowly expressed genes (expression value < 0.3) and then retaining top 500,000 significant co-expression pairs between genes (*FDR* < 0.01, Pearson correlation test) using gene expression data from adult human brain[55].

### Network analysis and visualization

To uncover disorder-specific topologies associated with BAG, subnetworks corresponding to each disorder and BAG-associated gene were extracted using the criterion that one side of the edge is a disorder risk gene or BAG-associated gene. The maximal connectivity subgraph was then extracted as the disorder-specific topology. The connectivity score (Cs) was used to estimate the strength of the connections within the molecular network, defined as follows:

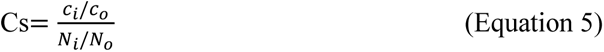

where *C_i_* and *C_o_* respectively represented the number of connections between a node with other nodes within a subset and the number of all the connections involving this node in the network, and *N_i_* and *N_o_* represented the number of nodes in the subset and the number of all nodes in the molecular network, *S_c_* more than one mean that the node has stronger connections to the inside than to the outside.

To estimate the correlation of two sub-networks, such as a BAG-associated sub-network and an ADHD-related sub-network, the Jaccard distance index was employed to measure the enrichment between the networks, defined as follows:

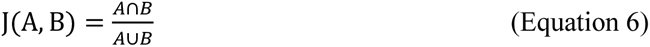

where A was the node set of BAG-associated sub-network, B was the node set of ADHD-related sub-network.

For better visualization, we created nodes for each BAG-associated gene and other risk genes, connecting all nodes via transcriptional regulation or protein-protein interaction. Finally, we arranged the networks using a perfuse circle layout. In the figure, we only presented the BAG-associated subnetwork. Network visualization was performed using Cytoscape[133].

## Ethical statement

Not applicable.

## Data availability

The original public datasets were available via the following URLs. IXI (https://brain-development.org/ixi-dataset/), SALD (http://fcon_1000.projects.nitrc.org/indi/retro/sald.html), CBIC (https://research.weill.cornell.edu/node/3598), AIBL (https://aibl.csiro.au/), OASIS1 (https://www.oasis-brains.org/), UK Biobank (https://www.ukbiobank.ac.uk/), ADNI (https://adni.loni.usc.edu/), and protein to protein interaction network (https://string-db.org/). The GWAS summary statistics dataset of BAG was uploaded to the GWAS Catalog (GCP001172).

## Code availability

The codes for this study can be found at Github (https://github.com/Soulnature/BrainAgepredict) and BioCode (https://ngdc.cncb.ac.cn/biocode/tool/BT007788).

## CRediT authorship contribution statement

**Xingzhong Zhao:** Methodology, Software, Formal analysis, Visualization, Writing – original draft, Writing – review & editing. **Anyi Yang:** Formal analysis. **Jing Ding:** Supervision, Writing – review & editing. **Yucheng T. Yang:** Conceptualization, Supervision, Writing – original draft, Writing – review & editing. **Xing-Ming Zhao:** Conceptualization, Supervision, Writing – original draft, Writing – review & editing. All authors have read and approved the final manuscript.

## Competing interest

The authors declare that they have no competing interests.

## Acknowledgements

The authors would like to thank the Institute of Science and Technology for Brain-Inspired Intelligence at Fudan University for supporting high-performance computing. The authors also want to thank Professor Jianfeng Feng for his contribution to accessing the original data sets. This work was partly supported by National Natural Science Foundation of China (T2225015, 61932008), Shanghai Science and Technology Commission Program (23JS1410100, 24JS2810100), Hainan Province Science and Technology Special Fund (ZDYF2024SHFZ058), National Key R&D Program of China (2023YFF1204800, 2020YFA0712403), Lingang Laboratory & National Key Laboratory of Human Factors Engineering Joint Grant (LG-TKN-202203-01), and Natural Science Foundation of Shanghai (21ZR1408100).

## Notes

### Competing Interest Statement

The authors have declared no competing interest.

